# Cohort Identification Using Semantic Web Technologies: Triplestores as Engines for Complex Computable Phenotyping

**DOI:** 10.1101/2021.12.02.21267186

**Authors:** Emily R. Pfaff, Robert Bradford, Marshall Clark, James P. Balhoff, Rujin Wang, John S. Preisser, Kellie Walters, Matthew E. Nielsen

## Abstract

**Background:** Computable phenotypes are increasingly important tools for patient cohort identification. As part of a study of risk of chronic opioid use after surgery, we used a Resource Description Framework (RDF) triplestore as our computable phenotyping platform, hypothesizing that the unique affordances of triplestores may aid in making complex computable phenotypes more interoperable and reproducible than traditional relational database queries.

To identify and model risk for new chronic opioid users post-surgery, we loaded several heterogeneous data sources into a Blazegraph triplestore: (1) electronic health record data; (2) claims data; (3) American Community Survey data; and (4) Centers for Disease Control Social Vulnerability Index, opioid prescription rate, and drug poisoning rate data. We then ran a series of queries to execute each of the rules in our “new chronic opioid user” phenotype definition to ultimately arrive at our qualifying cohort.

**Results:** Of the 4,163 patients in the denominator, our computable phenotype identified 248 patients as new chronic opioid users after their index surgical procedure. After validation against charts, 228 of the 248 were revealed to be true positive cases, giving our phenotype a PPV of 0.92.

**Conclusion:** We successfully used the triplestore to execute the new chronic opioid user phenotype logic, and in doing so noted some advantages of the triplestore in terms of schemalessness, interoperability, and reproducibility. Future work will use the triplestore to create the planned risk model and leverage the additional links with ontologies, and ontological reasoning.

## BACKGROUND

*Electronic health record (EHR)-based computable phenotypes* (hereafter “computable phenotypes”) are algorithms used to query “data captured in the delivery of healthcare… to identify individuals or populations (i.e., cohorts) with conditions or events relevant to” some use case.(1) An increasing number of clinical studies rely upon one or more computable phenotyping methods to define cohorts of interest.(2)

Despite its importance, computable phenotyping suffers from a number of inefficiencies and pitfalls. Most methods rely solely upon the data available in the EHR, which is imperfect, suffers from missingness, may not represent the “true” patient state, and can be semantically ambiguous.(1) Another major barrier to achieving robust, sharable, machine readable computable phenotypes is the way in which most healthcare institutions choose to structure and store their EHR data—in siloed, relational data warehouses, disconnected from other potentially relevant data sources and structured according to a custom or vendor-specific data model.

In this study, we describe our efforts to address some of these pitfalls as we executed a computable phenotype to identify previously opioid-naïve patients who became new chronic opioid users after a surgical procedure. This research question had complex data needs, with both an intricate phenotype requiring access to both EHR and claims data, as well as a need to link in community-level data for use in the eventual statistical model. Considering this complexity, we wanted to test whether this computable phenotype could be successfully run with SPARQL queries against a resource description framework (RDF) triplestore, rather than our standard method of running SQL queries against a relational database. In addition to determining whether the triplestore-driven phenotyping and data linkage approach was indeed feasible, we also sought to determine whether this approach is actually advantageous for certain complex phenotyping and linkage efforts.

### Computable Phenotyping

In the years following the 2009 HITECH Act, which incentivized and greatly increased usage of EHRs in American health care institutions, computable phenotyping has become a common mechanism to efficiently identify patient cohorts for clinical operations, quality improvement, and research purposes. The ideal computable phenotype is shareable, data model agnostic, semantically unambiguous, machine readable, and publicly accessible. This ideal has not yet been achieved at scale, but some version of this ideal has been advocated by many.(1,3–7)

However, there are many caveats to consider when using EHR data to define populations. The most essential point is that “the EHR is not a direct reflection of the patient and physiology, but a reflection of the recording process inherent in healthcare with noise and feedback loops.”(8) As such, important data can be missing or inaccurately recorded, the data may not reflect the full complexity of a patient’s condition, and the data may be biased toward high utilizers (i.e., sicker patients will generate more data).(4,8,9) Moreover, the context in which EHR data is collected, which can either be clinical or administrative, has a major impact on the meaning of the collected data.(1,10) EHR-driven computable phenotyping, then, requires acknowledgment and understanding of these caveats in balance with its power and efficiency as a method, and may benefit from the inclusion of complementary, non-EHR data sources (such as claims or community data) to fill in some of the gaps.

### Triplestores for Storing and Analyzing Clinical Data

A *triplestore* is the storage mechanism for data formatted in RDF. RDF is a W3C standard for data exchange on the web, represents data as a graph, and is heavily intertwined with the concept of the semantic web. Triplestores store data as triples, or statements in the format *subject – predicate – object*. (E.g., Michael Strauss (*subject*) is a (*predicate*) physician (*object*)). This restrictive format is at the same time quite flexible in its simplicity, as a wide variety of information can be encoded as triples. The rigidity of the triple format, however, is what allows the data contained in triples to be understood and consumed by different agents across the web—in other words, enabling interoperability. A number of prior studies have used triplestores for storing and analyzing EHR data, with a variety of rationales for doing so,(11–21) though this approach appears to exist outside the computable phenotyping mainstream. Some of the specific affordances of triplestores for clinical data are listed below.

### Linking with Ontologies and Terminologies

Triplestores are a natural fit for working with ontologies and terminologies, many of which are published in RDF or Web Ontology Language (OWL) format and can be easily ingested by a triplestore. An *ontology* is a systematic representation of knowledge; a way to consistently classify and define data across users, databases, institutions, and nations. *Terminologies* are controlled vocabularies in a given domain, such as ICD (the International Statistical Classification of Diseases and Related Health Problems) for diagnoses and procedures, or LOINC (Logical Observation Identifiers Names and Codes) for laboratory tests and results. Ontologies and terminologies provide common terms and definitions for, in this case, clinical concepts. Ontologies also allow for computational logical reasoning on the relationships between concepts, which is not the case with terminologies.(22)

As data get larger in volume and more heterogeneous in structure, use of ontologies and terminologies to label data is increasingly important to ascribe valid, consistent meaning to data points across disparate data sources. Moreover, mapping clinical data to an ontology helps to promote interoperability and complex querying based on logical reasoning.

### Schemalessness

*Schemalessness*, or tolerance of loosely modeled data, is a hallmark feature of triplestores and all “Not Only SQL,” or NoSQL, database technologies. Because clinical data is dynamic, changing frequently with new discoveries, new regulations, and advancements in EHRs, an adaptable data model is highly advantageous. Moreover, tolerance of heterogeneity makes it easier to link EHR data with external data sources (e.g., genomic data, geographically-based data, sensor data), which have their own structures distinct from that of the clinical data. In contrast, altering the data model of a relational database can come at a high cost, either requiring adding columns to existing tables (and creating sparsity within existing tuples) or adding new tables.

It is worth pointing out that having no data model at all would be as disadvantageous as having too rigid a model. With no model, data validity suffers (e.g., with three different ways to store blood pressure measurements, critical data could be easily missed in a query), and interoperability would be impossible. To achieve balance, one can use a standard clinical data model (e.g., openEHR, HL7, etc.) in tandem with a NoSQL database. Such models are specifically designed to accommodate changing requirements, and fit nicely within different NoSQL technologies. This approach allows a database to be technically schemaless, but with guardrails.

### Modeling Highly Related Data

Triplestores also excel at storing and querying “highly related” data. This includes hierarchical data, where one needs to retrieve, for example, the parent of the parent of the parent of a given concept. This may also include situations where the relationship between data is as important to model and expand upon as the data itself, such as a relationship between a gene and a disease that changes in the presence of certain drugs, environmental exposures, or other gene variants. Triplestores (and other graph databases) excel in this area specifically, due to their ability to materialize relationships as queryable concepts. Clinical data is relationship dense—diseases have causes, medications have side-effects, treatments have outcomes—and may be particularly well-suited to the graph structure for this reason.

Each of these affordances is a rationale behind our decision to use a triplestore to execute a particularly complex computable phenotype—one that required linkage of heterogeneous data sources and semantic harmonization across different datasets. Though our use case does not take full advantage of all of these capabilities, it serves as a starting point for more elaborate phenotyping use cases in the future.

## METHODS

The study motivating our triplestore-driven phenotyping approach aims to devise a model to determine a previously opioid-naïve patient’s risk of becoming a new chronic opioid user after a surgical procedure resulting in an opioid prescription for post-operative pain. Before beginning work on the risk model, it was first necessary to define the computable phenotype to identify existing postsurgical new chronic opioid users, whose shared characteristics would enable us to define the risk model for future patients.

The data sources used for both the phenotyping and the risk modeling effort included (1) UNC Health Care System EHR data; (2) claims data from a major commercial insurer in the Southeast; (3) American Community Survey (ACS) data (2012-2016);(23) and (4) CDC Social Vulnerability Index (SVI),(24) opioid prescription rate,(25) and drug poisoning rate data (all 2016).(26)

### Phenotype Definition

The criteria used to define new postsurgical chronic opioid users (adapted by author MN based on a prior study(27)) depends upon first applying criteria to the base population of surgical patients so that those with recent history of opioid use or prior opioid use disorder are excluded. These criteria are:

- Patient has one CPT (procedure) code in the range 10000 – 69990 (the “surgery” range) between 4/14/2014 and 3/31/2017.
- Patient does NOT have a second CPT code in the surgery range within six months (after) the index surgery.
- Patient has at least one opioid prescription between 30 days prior to the index surgery and 14 days after the index surgery.
- Patient does NOT have an opioid prescription between 1 year prior and 30 days prior to the index surgery (i.e., the patient was “opioid naïve” before surgery).
- Patient does not have a diagnosis code for “opioid use disorder” (ICD-9: 304.0X, 304.7X; ICD-10: F11.X) prior to the index surgery.
- Patient had insurance with the commercial insurer from whom we obtained claims data at the time of the index surgery. (Not relevant to the patient’s status as a chronic opioid user, but necessary in this study to ensure linkage with available claims data.)

Among the resulting set of previously opioid-naïve patients, we define new chronic opioid users as:

- Patient has at least one opioid prescription between 90 and 180 days after the index surgery (suggesting that they are still using opioids well after the surgery at which they were originally prescribed).

This rule-based computable phenotype is possible to implement with SQL code against EHR data— however, stopping at this single source of data would result in a skewed picture. This is because medication data is such an essential part of this phenotype—and as such, we care not only about medications that were prescribed by UNC physicians, but any medications that the patient may have received, from anywhere. In order to make use of non-UNC medication data, we needed to incorporate insurance claims data, which allowed us to work with a superset of patient medication data—all medications billed to the patient’s insurance, regardless of prescriber. Merging claims-based medication data with EHR-based medication data introduced one level of structural heterogeneity.

To add further heterogeneity, we also recognized the need to incorporate community-level data sources in order to access socioeconomic data about the cohort for use in the risk model. While publicly available, these data sources are not available within UNC’s clinical data warehouse, and would have to be downloaded and modeled appropriately for incorporation with the EHR and claims data.

Considering the challenges of incorporating these disparate data sources for unified analysis, as well as the affordances described earlier, our team decided on a triplestore approach.

### Data Transformation Pipeline

As the three data sources are highly heterogeneous, we developed a transformation pipeline to take each data source from its raw form to RDF. This pipeline is illustrated in Figure 1, and described in detail in the text below.

**Figure 1.**
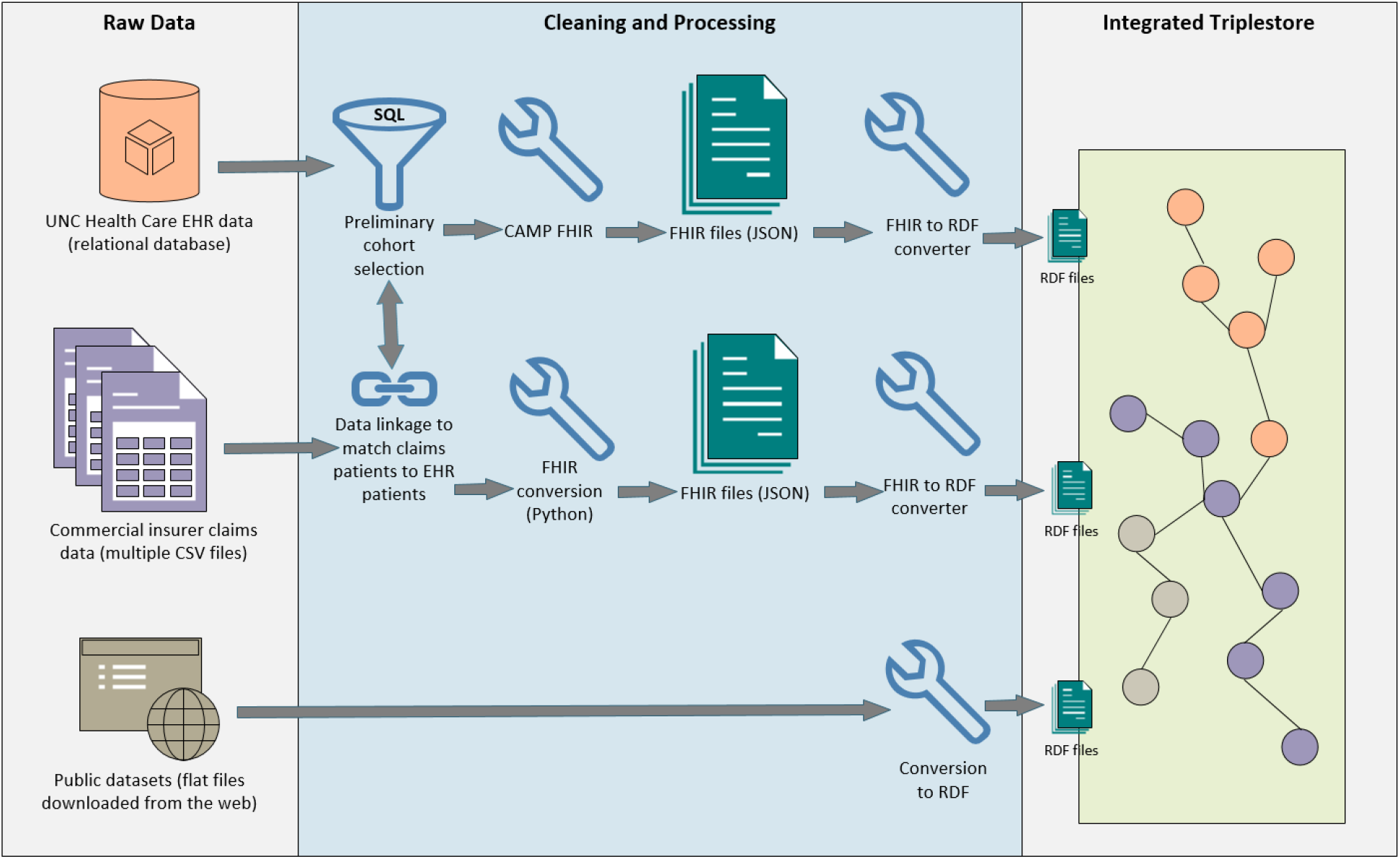
Data transformation pipeline to take EHR, claims, and geographically based datasets from their native formats to RDF, and integrate them in a Blazegraph triplestore.

### EHR Data

UNC’s EHR data is sourced from the Carolina Data Warehouse for Health (CDWH), UNC Health’s enterprise clinical data warehouse. An analyst was tasked with extracting a cohort meeting the specifications of the rule-based computable phenotype listed above, with the exception of the last rule. (As we wanted to compare patients who did and did not become new chronic opioid users for the risk model, we did not want to limit the cohort only to new chronic users at this point.) Because of the limitations of the EHR data, the resulting cohort was known to contain false positives (i.e., patients who qualified for the cohort using EHR data only, but would later be excluded based on claims data).

We extracted EHR data for this cohort in the domains of patient demographics, encounter details, diagnoses, procedures, medications, smoking status, and pain scores. To convert the extracted data to HL7 Fast Healthcare Interoperability Resources (FHIR) format (version 3.0.1) we used CAMP FHIR,(28,29) an open-source data conversion utility built to convert EHR data to FHIR format. Once the data were in FHIR format, we were able to use an open-source FHIR-to-RDF conversion utility(30) to output the data in RDF and ready it for loading into the triplestore.

### Claims Data

We produced a finder file containing identifiers for all of the patients identified from the EHR data, which an analyst used to perform a deterministic linkage with the claims cohort. Matching claims data were extracted in the domains of plan membership details, medications, diagnoses, and procedures. The latter three domains were converted to FHIR format using a Python script, and then converted to RDF using the same conversion utility as the EHR data. The plan membership data were manually converted to RDF using R,(31) as it did not fit cleanly within the FHIR specification.

### Community Data

Though non-clinical data was not required to run the computable phenotype to identify existing chronic users, the research team did want to include socioeconomic and other geographically-based factors in the eventual risk model. The chosen datasets (ACS and CDC SVI, opioid prescribing rate, and drug poisoning rate data) are all publicly available, but do not have a unified structure. For the purpose of querying these data within our triplestore, we minted custom URIs to represent the predicates required to represent the structure of the data (e.g., *livesInTract*).

### Other Data

Four additional datasets were loaded in order to aid querying and make connections between datasets in the triplestore. First was a patient-to-census-tract crosswalk, extracted from the EHR. Second, we loaded a census-tract-to-county crosswalk, to enable linkage with both tract-based data (e.g., ACS) and county-based data (e.g., CDC opioid prescribing rates).

Third, in order to consistently query medication data across EHR and claims data, we loaded a crosswalk between National Drug Codes (NDC) and RxNorm codes. Claims data uses NDC codes, which are representative of the exact medication dispensed by a pharmacy for a submitted claim, while UNC’s EHR medications are RxNorm coded. Using FHIR, it is possible to tie multiple codes to a single Medication resource where they share a common meaning. To accomplish this, we used a Python program called SMOREs(32) to translate between NDC and RxNorm and stored both values within the same Medication resource that was subsequently converted to RDF.

Finally, we loaded a subset of the Department of Veterans Affairs National Drug File (VANDF), crosswalked to RxNorm codes. This was loaded to make use of VANDF’s categorization scheme for medications in our queries, allowing us to query by high-level medication categories (opioids, benzodiazepines) rather than matching on hundreds of individual RxNorm codes.

After each data source (EHR, claims, community, other) was converted to RDF, we loaded each RDF file into a Blazegraph(33) triplestore.

### Phenotyping with SPARQL

We wrote a series of SPARQL queries to execute the same rule-based logic as the original SQL queries that extracted the EHR-based cohort. Because the data were now integrated, running the same logic a second time could (and did) exclude patients who had originally qualified for the EHR cohort. Examples of such situations included patients who didn’t have a disqualifying pre-surgery opioid prescription in UNC’s EHR data, but did have one (or more) reflected in the claims data, and patients who only had one surgical procedure in a six-month period in UNC’s EHR data, but whose claims data showed an additional (disqualifying) procedure at another institution.

A final SPARQL query was written to implement the final rule of the phenotype (the presence of an opioid prescription between 90 and 180 days after the index surgery), which would define our outcome variable for each patient in the cohort: CHRONIC_USER_YN.

## RESULTS

The number of triples loaded into Blazegraph from each data domain are listed in Table 1.

**Table 1.** All data domains included in the Blazegraph triplestore, with number of triples.

| **Data Source** | **Data Domain** | **Format** | **# of Triples Loaded** |
| --- | --- | --- | --- |
| **EHR** | Patient | FHIR | 341,248 |
|  | Encounter | FHIR | 18,180,947 |
|  | Procedure | FHIR | 10,158,248 |
|  | MedicationRequest | FHIR | 80,017,422 |
|  | Medication | FHIR | 549,175 |
|  | Observation (vitals & smoking status) | FHIR | 57,412,289 |
|  | Condition | FHIR | 100,614,527 |
| **Claims** | Plan membership data | Custom | 12,417 |
|  | MedicationDispense | FHIR | 22,176,831 |
|  | Medication | FHIR | 629,928 |
|  | Procedure | FHIR | 16,483,352 |
|  | Condition | FHIR | 12,155,855 |
| **Geographically Based** | CDC SVI* | Custom | 256,464 |
|  | CDC opioid prescribing rates* | Custom | 98 |
|  | CDC drug poisoning rates* | Custom | 200 |
|  | American Community Survey*† | Custom | 2,265,240 |
| **Other** | Census tract to county crosswalk* | Custom | 2,195 |
|  | Patient to census tract crosswalk | Custom | 4,157 |
|  | Department of Veterans Affairs<br>National Drug File (opioid and<br>benzodiazepine categories only) to<br>RxNorm crosswalk | Custom | 1,438 |

\* Despite national information being available, only North Carolina data was loaded for the purposes of this study.
† Only a subset of the available American Community Survey variables were loaded for the purposes of this study.

We initially loaded 15,954 patients that qualified as previously opioid-naïve patients with a qualifying surgical procedure, executed against EHR data only. Once the matching claims data were loaded and the computable phenotype (except for the final rule) was re-executed over the integrated data using SPARQL, the number of qualifying patients dropped to 4,163. The reasons for this drop are illustrated in Figure 2.

**Figure 2.**
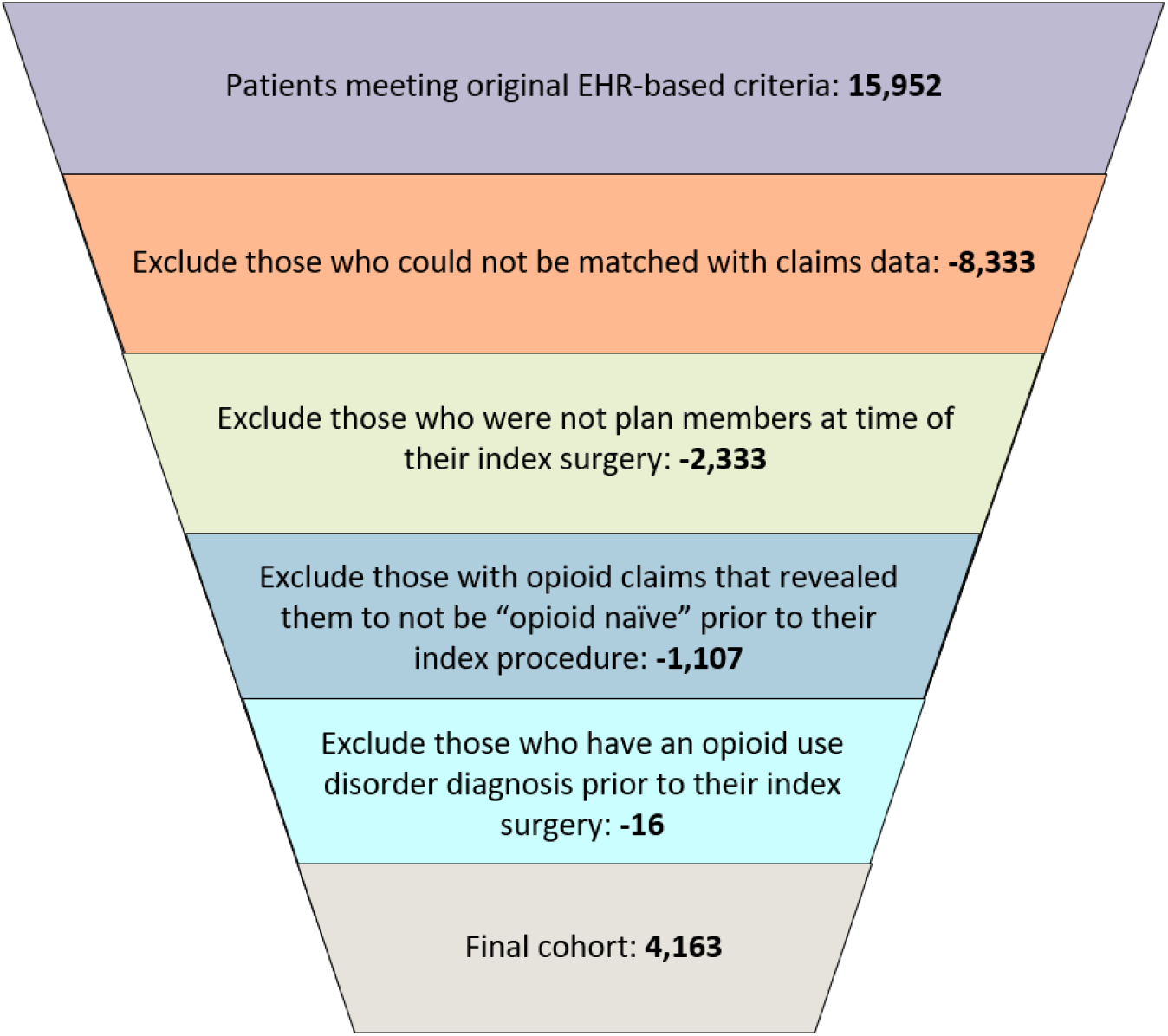
Illustration of cohort narrowing as phenotype rules were applied in the triplestore.

Of the 4,163 patients in the final cohort, 248 patients were identified as new chronic opioid users after their index surgical procedure. Patient identifiers for this group were given to two clinicians to manually chart review in order to validate the actual presence of new chronic opioid use in the patient’s record. Of the 248 patients identified by the phenotype, 228 were found to be true new chronic users through chart review. The remaining 20 patients qualified for the computable phenotype, but upon review of their charts, the procedure defined as their index surgery (which qualified them for the cohort) appeared not to have truly been a “surgery,” despite being coded in the surgical range. One possible explanation for this is that the CPT “surgery” range of 10000 through 69900 is very broad, and may have resulted in the computable phenotype considering certain events as “surgeries” when a clinician would not see them as such. Based on these results, this computable phenotype has a positive predictive value of 0.92 for identifying new chronic opioid users after surgery.

Of the 248 patients identified as new chronic opioid users, 16 were identified by EHR data alone, 39 were identified by both EHR and claims data, and 193 were identified by claims data alone. (“Alone,” in this case, means that while the patient had data in both the EHR and claims, the opioid prescription(s) that qualified them as a new chronic user between 90 and 180 days from their index surgery] was only present in one of the two sources.)

4,121 of the 4,163 patients were able to be linked with a census tract and county, based on their address recorded in the EHR as of March 31, 2017 (the end of the study period). This enables us to connect these patients with a selection of socioeconomic variables (including Social Vulnerability Index, drug poisoning rate, and unemployment rate) for potential inclusion in the new chronic opioid user risk model.

## DISCUSSION

### Primary Results

Our main finding is that triplestores are an effective platform for complex computable phenotyping, enabling cross-querying among heterogeneous datasets and offering some functionality beyond that of a relational database. For this particular phenotyping project, combining claims data with EHR data was particularly advantageous, allowing us to identify many more patients qualifying for the phenotype than using EHR data alone. Converting the EHR data and the claims data to a common format (FHIR, then RDF) and loading it in a triplestore made integration and cross-querying very straightforward, essentially like querying a single source of data.

It is worth noting that for a dataset of this size and scope, use of this triplestore-driven approach was not a requirement. Rather, we view this work as a successful proof of concept for future work (detailed below) involving phenotypes that require (or could benefit from) the inclusion of much larger and varied ontologies and datasets (e.g., the Human Phenotype Ontology, the entirety of SNOMED CT, the Gene Ontology, etc.).

### Advantages of the Triplestore Approach

#### Schemalessness

The triplestore’s schemalessness proved particularly valuable in the context of the community datasets. Beyond converting from CSV to RDF format, almost no transformation was needed to prepare these data sources for ingestion in the triplestore. Should we have found additional publicly available data sources that we wanted to use, it would have taken very little effort to incorporate them. This capability holds a lot of promise for future studies, particularly due to the recognition of the importance of social determinants of health variables, which are often absent in EHR and claims data, in clinical research.(34–38)

#### Interoperability and Reproducibility

The FHIR representation standard is becoming increasingly supported by major EHR vendors—in effect, forcing institutions to standardize their data. In addition to defining specific resources and fields, FHIR enforces the use of established code sets (e.g., LOINC, SNOMED CT, ICD-9/-10) or FHIR-specific value sets in many of its fields in order to maximize standardization. However, as FHIR data exists in flat files (JSON- or XML-formatted), it is not inherently “queryable” unless additional processing steps are taken. In this case, loading FHIR data in a triplestore enables its use for computable phenotyping.

The interoperability benefits of FHIR, compared with the interoperability (by definition) of semantic web technologies, enabled us to create a phenotype that should be transferrable to any other site with few local changes, so long as the institution can output EHR data in FHIR format. (This particular study offered a proof of this concept, in that FHIR enabled us to make EHR and claims data interoperable despite their native formats being completely different.) As FHIR moves toward becoming a *lingua franca* for EHR data, FHIR-based phenotypes such may gain reproducibility advantages over phenotypes that run against institution-, data network- or vendor-specific data models.

### Limitations and Lessons Learned

Because the FHIR format creates an enormous number of triples (as shown in Table 1), it would seem to be a best practice to narrow down the initial cohort as much as possible before transformation to RDF. It would be less feasible to try to persist an entire clinical data warehouse in RDF. Similarly, within a given data domain (particularly a large one, like Condition), it would be more efficient to pare down the set of diagnoses to only those needed for the given research question prior to loading. With that said, if a study’s objective is less targeted and more exploratory, this may not be desirable. For such a use case, sufficient resources to load and compute over all those triples would be the primary concern.

Additionally, as with any use case that involves data transformation, there is always a chance that information can be lost over the course of the transformation pipeline. For this reason, we took the time to validate our final cohort against the source data system (the EHR), and would advocate doing so for future cohorts identified in this way.

### Future Work

Now that the computable phenotype has been executed, statistical analysis has commenced in order to establish a risk model that can be applied to patients *de novo*. This work will occur outside of the triplestore. In parallel with this effort, we identified several new directions to take the triplestore phenotyping methodology that will make this framework valuable for future use cases.

We see great potential in leveraging additional ontologies (other than the medication ontologies used in this study) in future work. Ontologies allow users to work with information at both the “data level” and the “knowledge level.”(13) As data at the knowledge level is not often stored in the EHR itself, the ability to link to external sources containing that knowledge adds richness to a clinical data store.(15,18,39)

“OWL reasoning,” another capability afforded by RDF, is an extension of this knowledge level, in that new assertions can be concluded based on existing assertions in the triplestore.(21,40,41) For this project specifically, we used a small subset of VANDF to query medications at the knowledge level to enable easier querying (e.g., querying by rolled-up categories rather than hundreds of individual medication codes). With this success in mind, we believe that linking with ontologies and leveraging OWL reasoning could potentially reveal new hypotheses to explore, via patterns in the data that may be hidden at the individual medication code level.

Use of standard ontologies also can enable linkage with other external datasets, in addition to the community data we used here. Additional external data sources may include one or more of the datasets available on the web as Linked Open Data, or LOD, an open data repository expressed in triples. Many of the datasets available as LOD are biomedical in nature, and could allow one to link EHR data with public data to discover, for example, gene-disease associations, genotype-phenotype associations, or drug-drug interactions.(11,12,14,18)

## CONCLUSION

Though computable phenotypes are written, executed, and shared with great frequency today, computable phenotyping as a discipline has not yet reached the ideal of consistently producing phenotypes that are interoperable, reproducible, and capable of integrating heterogeneous data sources. Some of these challenges may be addressed by exploring alternatives to siloed, relational data warehouses for storage and analysis of clinical data. Semantic web technologies may be such an alternative.

## Data Availability

The electronic health record and claims data analyzed in this study are not publicly available due to restrictions of the Health Insurance Portability and Accountability Act (HIPAA). American Community Survey data (2012-2016) is available from the US Census Bureau at https://www.census.gov/acs/www/data/data-tables-and-tools/data-profiles/2016. Social vulnerability index data (2016) are available from the Centers for Disease Control at https://svi.cdc.gov/data-and-tools-download.html. US county opioid prescribing and drug poisoning rate data (2016) are available from the Centers for Disease Control at https://www.cdc.gov/drugoverdose/maps/rxcounty2016.html and https://data.cdc.gov/NCHS/NCHS-Drug-Poisoning-Mortality-by-County-United-Sta/pbkm-d27e, respectively.

https://www.cdc.gov/drugoverdose/maps/rxcounty2016.html

https://svi.cdc.gov/data-and-tools-download.html

https://data.cdc.gov/NCHS/NCHS-Drug-Poisoning-Mortality-by-County-United-Sta/pbkm-d27e

## LIST OF ABBREVIATIONS

ACS: American Community Survey
CDC: Centers for Disease Control and Prevention
CDWH: Carolina Data Warehouse for Health
CPT: Current Procedural Terminology
EHR: Electronic health record(s)
FHIR: Fast Healthcare Interoperability Resources
HITECH Act: Health Information Technology for Economic and Clinical Health Act
HL7: Health Level 7
ICD: International Statistical Classification of Diseases and Related Health Problems
JSON: JavaScript Object Notation
LOD: Linked Open Data
LOINC: Logical Observation Identifiers Names and Codes
NDC: National Drug Code
NoSQL: Not only SQL
OWL: Web Ontology Language
PPV: Positive predictive value
RDF: Resource Description Framework
SPARQL: SPARQL Protocol and RDF Query Language
SQL: Structured query language
SVI: Social Vulnerability Index
UNC: University of North Carolina at Chapel Hill
URI: Uniform resource identifier
VANDF: Department of Veterans Affairs National Drug File
W3C: World Wide Web Consortium
XML: Extensible Markup Language

## DECLARATIONS

### Ethics

The work described was approved by the Institutional Review Board at UNC Chapel Hill (#18-1154).

### Consent to Publication

Not applicable.

### Author contributions

E.R.P. drafted the initial manuscript. M.E.N. defined the phenotype and provided clinical expertise during analysts. E.R.P., R.B., and M.C. performed data cleaning, transformation, and analysis. J.P.B. provided ontological and technical expertise during analysis. R.W. and J.S.P. provided statistical consulting and analysis. K.W. selected appropriate SES variables and performed literature review. All authors edited the manuscript draft and approved the final content.

## Acknowledgements

The authors thank Charan Mohan, Mark Ehlers, and Nessim Abu-Saif for performing manual chart reviews to assess phenotype performance, Adam Lee for serving as an EHR honest broker, Lily Wang for serving as a claims honest broker, and James Champion for his assistance with implementing CAMP FHIR as part of our data transformation pipeline.

